# Differences Between Reported COVID-19 Deaths and Estimated Excess Deaths in Counties Across the United States, March 2020 to February 2022

**DOI:** 10.1101/2023.01.16.23284633

**Authors:** Eugenio Paglino, Dielle J. Lundberg, Zhenwei Zhou, Joe A. Wasserman, Rafeya Raquib, Katherine Hempstead, Samuel H. Preston, Irma T. Elo, Andrew C. Stokes

**Affiliations:** Department of Sociology and Population Studies Center, University of Pennsylvania, Philadelphia, PA; Department of Global Health, Boston University School of Public Health, Boston, MA; Department of Health Systems and Population Health, University of Washington School of Public Health, Seattle, WA; RTI International, Research Triangle Park, NC; Robert Wood Johnson Foundation, Princeton, NJ

**Author notes:** CORRESPONDING AUTHOR: Andrew C. Stokes, 801 Massachusetts Avenue, Crosstown Building 362, Boston, MA, 02118.

## Abstract

Accurate and timely tracking of COVID-19 deaths is essential to a well-functioning public health surveillance system. The extent to which official COVID-19 death tallies have captured the true toll of the pandemic in the United States is unknown. In the current study, we develop a Bayesian hierarchical model to estimate monthly excess mortality in each county over the first two years of the pandemic and compare these estimates to the number of deaths officially attributed to Covid-19 on death certificates. Overall, we estimated that 268,176 excess deaths were not reported as Covid-19 deaths during the first two years of the Covid-19 pandemic, which represented 23.7% of all excess deaths that occurred. Differences between excess deaths and reported COVID-19 deaths were substantial in both the first and second year of the pandemic. Excess deaths were less likely to be reported as COVID-19 deaths in the Mountain division, in the South, and in nonmetro counties. The number of excess deaths exceeded COVID-19 deaths in all Census divisions except for the New England and Middle Atlantic divisions where there were more COVID-19 deaths than excess deaths in large metro areas and medium or small metro areas. Increases in excess deaths not assigned to COVID-19 followed similar patterns over time to increases in reported COVID-19 deaths and typically preceded or occurred concurrently with increases in reported COVID-19 deaths. Estimates from this study can be used to inform targeting of resources to areas in which the true toll of the COVID-19 pandemic has been underestimated.

## Introduction

Excess mortality is a measure that has been widely used to assess the mortality impact of the COVID-19 pandemic.^1^ Excess mortality refers to the difference between the observed number of deaths that occurred during a given period and the number of deaths that would be expected based on mortality trends prior to the period.^2^ Prior estimates reveal that more than 1.1 million excess deaths occurred in the United States during the first two years of the pandemic, with approximately 620,000 occurring in the first year and 540,000 occurring in the second year.^3^ Most studies have found that the number of excess deaths substantially exceeded the number of deaths assigned to COVID-19 on death certificates.^4–8^ While a large body of research has investigated the spatial and temporal patterning of deaths assigned to COVID-19 and excess deaths during the pandemic,^4,5,9–11^ less attention has been given to how excess deaths not assigned to COVID-19 have been dispersed across the U.S..

Excess deaths not assigned to COVID-19 are deaths above those expected based on pre-pandemic trends that were not attributed to COVID-19 on death certificates.^12,13^ There are several reasons why excess deaths may not be assigned to COVID-19. First, the death may have been directly related to COVID-19 but unrecognized as a COVID-19 death by the certifier.^14^ This could occur due to a lack of COVID-19 testing before death, a lack of post-mortem COVID-19 testing due to limited death investigation resources, or resistance to assigning the death to COVID-19 from the deceased’s family or the death certifier as a result of personal or political beliefs.^15,16^ It could also relate to difficulty with assignment resulting from atypical presentations of COVID-19 symptoms or the presence of multiple comorbidities.^17,18^ Second, the death could be indirectly related to the pandemic as a result of health care interruptions or delays in health care that occurred especially during periods when COVID-19 cases increased and hospitals were overcrowded with COVID-19 patients.^19,20^ Lastly, the death could be indirectly related to the pandemic through the social and economic consequences of the pandemic.^21^ These deaths could be caused by changes in health resulting from food insecurity, housing instability, and other stressors and by increases in poisonings, suicide, and accidents.^22–26^

Identifying the spatial and temporal patterning of excess deaths not assigned to COVID-19 throughout the pandemic has important implications for the death investigation system and for social programs providing pandemic relief.^15^ Communities with high proportions of these deaths may represent areas where COVID-19 deaths were not accurately recognized because of flawed death investigation practices.^16^ These communities may be potential targets for additional training on death investigation.^27^ One important social program that has been established during the pandemic to provide economic relief to families who have lost loved ones to the pandemic is FEMA’s funeral assistance program. This program provides families with a maximum of $9,000 per deceased individual and a maximum of $35,500 per application to cover funeral costs.^28^ The application, however, requires that families provide a death certificate stating that the death may have been caused by or was likely a result of COVID-19. Deaths that were not assigned to COVID-19 on death certificates are not eligible for this program. As a result, communities with large numbers of these deaths may represent areas where this social program has had less impact. Thus, estimates of excess deaths not assigned to COVID-19 may help inform reforms to FEMA’s program and the development of new relief programs.

In the present study, we compare monthly estimates of excess mortality generated using a Bayesian hierarchical model based on all-cause mortality to county-month data on deaths assigned to COVID-19 for 3,127 counties during the first two years of the pandemic (March 2020 to February 2022). This comparison produces estimates of excess deaths not assigned to COVID-19 for each county-month. We then explore spatial and temporal patterning of these deaths relative to expected all-cause deaths across Census divisions and large metros, medium/small metros, and nonmetro areas throughout the first and second year of the pandemic.

## Data and Methods

### Experimental Design

This study focuses on the comparison between estimated excess mortality for the period March 2020 - February 2022 and official COVID-19 mortality. We modeled monthly all-cause excess mortality using a Bayesian hierarchical model for 3,127 counties for the period March 2020 to February 2022. We used publicly available data for the period January 2015 - December 2019 to fit the model, and examined a set of metrics to assess the goodness of fit and the model’s sensitivity to different specifications. Starting from the county-month estimates, we built geographically (Census divisions, metropolitan-nonmetropolitan categories) aggregated counts to investigate how excess mortality compared to COVID-19 mortality across the U.S. during the pandemic’s first two years.

### Data

We extracted monthly death counts at the county-level from the CDC WONDER online tool. See **Supplementary Text** for further details about data extraction procedures. We extracted all-cause death counts and COVID-19 death counts from the Multiple Cause of Death database using the provisional counts for 2021 and 2022 and the final counts for 2015-2020. We define COVID-19 deaths as those in which COVID-19 was listed as the underlying cause of death using the International Statistical Classification of Diseases and Related Health Problems, Tenth Revision (ICD-10) code *U07.1*. COVID-19 is listed as the underlying cause of death in approximately 92% of cases in which it is mentioned somewhere on the death certificate^29^. To transform the number of deaths into rates, we used publicly available yearly county-level population estimates from the Census Bureau (2010-2020^30^ and 2021^31^). To obtain monthly population estimates, we assumed the population grew linearly between each two time points. For the August 2021 - February 2022 period, for which no population estimates are available, we projected county-level population by computing county-specific average monthly growth rates for the period January 2018 - July 2021 (the most recent month for which Census Bureau estimates were available) and then applying these rates to the July 2021 population.

In some of the tables and figures, we present aggregate quantities by grouping counties into 3 metropolitan-nonmetropolitan categories (large metro, medium or small metro, and nonmetro) based on the 2013 NCHS Rural-Urban Classification Scheme for Counties.^32^ We also grouped counties into 9 Census Bureau Divisions (New England, Middle Atlantic, East North Central, West North Central, South Atlantic, East South Central, West South Central, Mountain, and Pacific). Finally, in some analyses, we stratified the Census Divisions by the metropolitan-nonmetropolitan categories. The **Supplementary Text** provides further details about the geographic classifications used in this study.

### Statistical Methods

We used a Bayesian hierarchical model to predict the monthly county-level number of deaths. Our methods are described in more detail elsewhere^3^, but in summary we started from the framework described in a prior paper^33^ and made some adaptations to fit our specific application. Let *y*_*ts*_ be the number of deaths in spatial unit *s* at time *t*. Let *P*_*ts*_ be the population of spatial unit *s* at time *t*. We assume a Poisson distribution for the number of monthly deaths *y*_*ts*_ and model the risk *r*_*ts*_ of dying using the following specification:

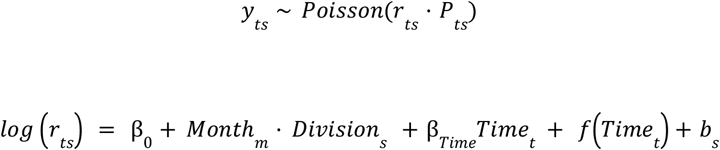

where β_0_ is the global intercept. We included dummy variables (fixed effects) for each combination of month and Census division to capture seasonal effects (allowing for division-level heterogeneity). The model incorporates a linear effect (captured by β_*Time*_) and non-linear effect *f*(·) of time (in months) since the start of the period (*t* = 1, 2, … with time 1 corresponding to January 2015). We added a non-linear effect of time, modeled as a first-order autoregressive process (AR1) model:

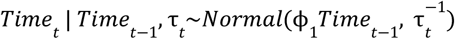

To capture the spatial structure of the data, we modeled county-level intercepts using the modified Besag, York and Mollie spatial model proposed by Riebler et al. (BYM2 model).^34^ This model is the sum of two random effects, a spatially unstructured one, 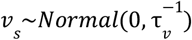 and spatially structured one *u*_*s*_. The term *b*_*s*_ in the model is thus defined as:

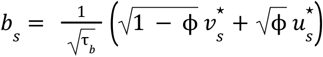

where 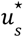 and 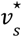 are standardized versions of *u*_*s*_ and *v*_*s*_ to have variance equal to 1. The term 0 ≤ ϕ ≤ 1 is a mixing parameter which measures the proportion of the marginal variance explained by the spatially structured effect.

We used minimally informative prior distributions for the fixed effects β_0_, the month-division specific intercepts *Month*_*m*_ · *Division*_*s*_ *m* = 1, 2, …, 12, the linear time effect β_*Time*_, and the ϕ_1_ parameter for the AR1 process. For the hyperparameters of the BYM2 model, ϕ and τ_*b*_, we adopted priors that tend to regularize inference while not providing too strong information, the so-called penalized complexity (PC) priors introduced by Simpson et al.^35^ In particular, for the standard deviation 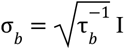 select a prior so that *Pr* (σ_*b*_ > 1) = 0. 01, implying that it is unlikely to have a spatial relative risk higher than *exp*(2) based solely on spatial or temporal variation. For ϕ we set Pr *Pr* (ϕ < 0. 5) = 0. 5 reflecting our lack of knowledge about which spatial component, the unstructured or structured one, should dominate the spatial term *b*_*s*_.

Finally, we also adopt PC priors for the remaining standard deviation 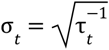 such that *Pr* (σ_*t*_ > 1) = 0. 01.

We fit the models using the Integrated Nested Laplace Approximation (INLA) method, through the R-INLA software package.^36^

To compare measures of excess mortality and differences between excess mortality and COVID-19 mortality across counties with different populations and numbers of deaths, we used relative measures of excess mortality throughout this study. Relative excess mortality refers to the number of excess deaths divided by the number of expected deaths in an area. Relative excess mortality not assigned to COVID-19 refers to the difference between excess deaths and deaths assigned to COVID-19 divided by the number of expected deaths in an area.

This study used de-identified publicly available data and was exempted from review by the Boston University Medical Center Institutional Review Board. Programming code was developed using R, version 4.1.0 (R Project for Statistical Computing) and Python, version 3.7.13 (Python Software Foundation).

## Results

**Table 1** presents estimates of excess deaths not assigned to COVID-19 in the U.S. and across Census divisions and metro-nonmetro categories. We estimated that between March 2020 and February 2022 there were 1,134,364 excess deaths (95% prediction interval (PI): 996,869 to 1,278,540) occurred in the U.S., of which 866,187 were assigned to COVID-19 and 268,176 (95% PI: 130,682 to 412,353) were not assigned to COVID-19. Proportionally, this indicates that 23.7% (95% PI: 11.5% to 36.4%) of excess deaths in the U.S. were not assigned to COVID-19. **Figure 1** shows the ratio of excess mortality to COVID-19 mortality in counties across the U.S., revealing significant heterogeneity in the proportion of excess deaths not assigned to COVID-19 throughout the country. The **Supplemental Table** shows this heterogeneity at the state-level.

**Table 1.**
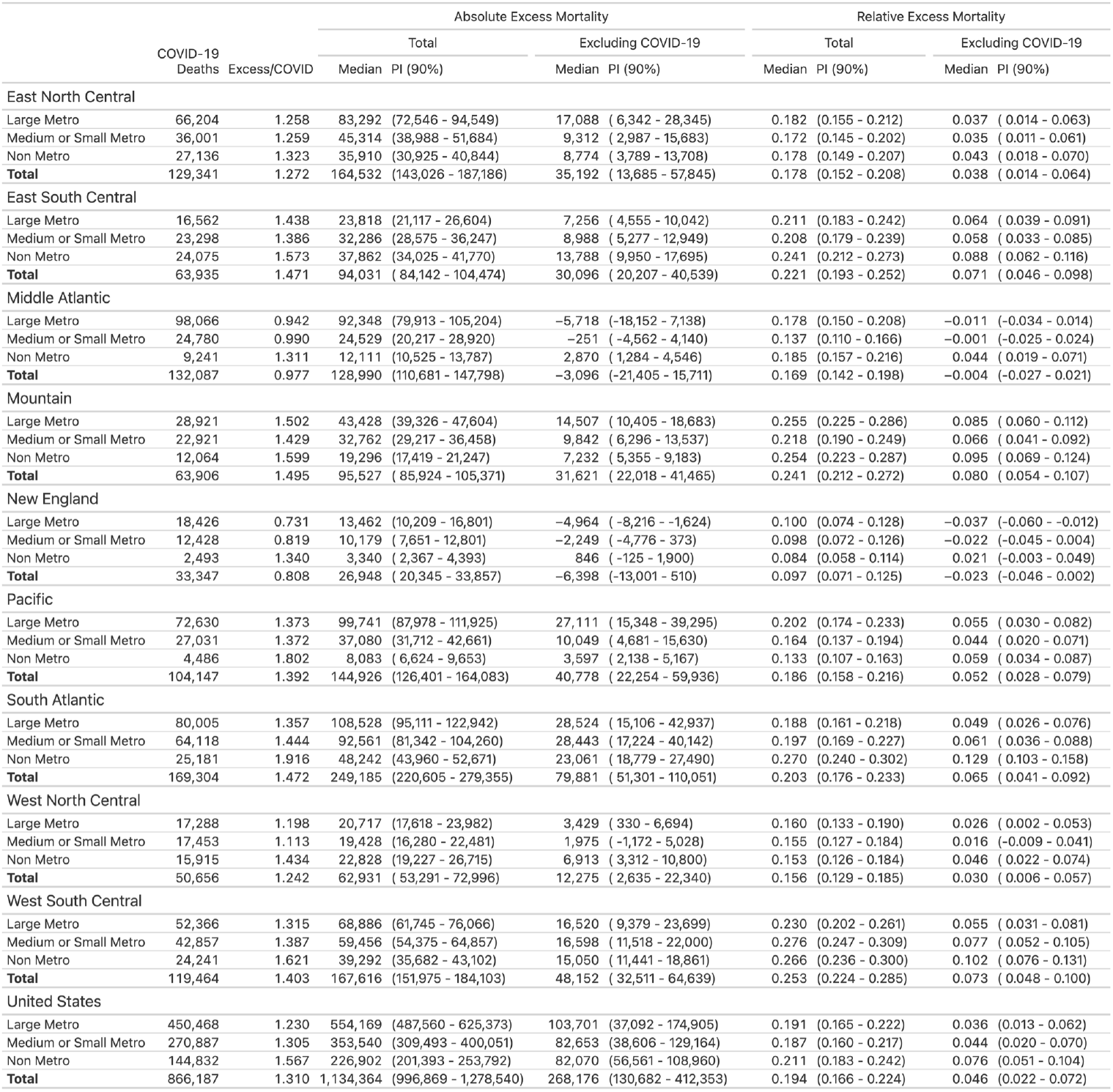
Excess mortality across U.S. Census Divisions and metro/nonmetro areas over the first two years of the pandemic.

**Figure 1.**
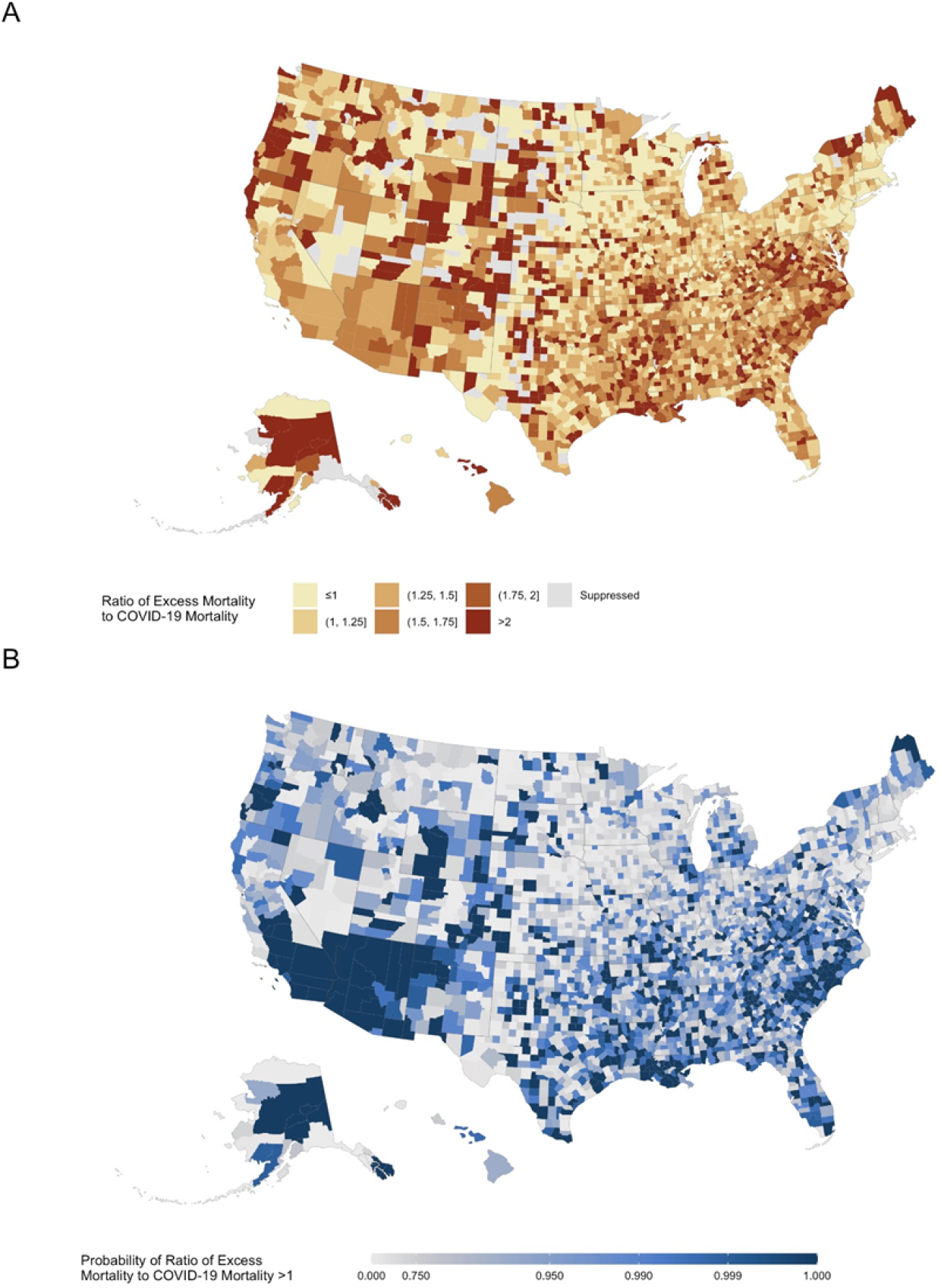
Ratio of excess mortality to COVID-19 mortality across U.S. counties, March 2020 - February 2022 <u>Notes</u>: Each county in the map is colored according to its ratio of excess mortality to COVID-19 mortality. Suppressed values reflect counties in which the cumulative number of COVID-19 deaths from March 2020 through February 2022 was less than 10 deaths.

### Estimates Across Metro-Nonmetro Categories

After subtracting deaths assigned to COVID-19, the total of deaths observed from March 2020 to February 2022 was 4.6% (95% PI: 2.2% to 7.2%) higher than expected based on pre-pandemic trends. This estimate also known as relative excess mortality for deaths not assigned to COVID-19 was higher in non-metro counties (7.6% [95% PI: 5.1% to 10.4%]) compared to medium or small metro counties (4.4% [95% PI: 2.0% to 7.0%]) and large metro counties (3.6% [95% PI: 1.3% to 6.2%]).

### Estimates Across Census Divisions

**Figure 2** compares excess deaths to COVID-19 deaths in counties across Census divisions, revealing that all divisions except the New England and Middle Atlantic divisions appeared to have more excess deaths than COVID-19 deaths. Relative excess mortality for deaths not assigned to COVID-19 was highest in the Mountain division and in the South. It was 8.0% (95% PI: 5.4% to 10.7) in the Mountain division, 7.3% (95% PI: 4.8% to 10.0%) in the West South Central division, 7.1% (95% PI: 4.6% to 9.8%) in the East South Central division, and 6.5% (95% PI: 4.1% to 9.2%) in the South Atlantic division. In contrast, relative excess mortality for deaths not assigned to COVID-19 was negative in the New England division (−2.3% [95% PI: -4.6% to 0.2%]) and in the Middle Atlantic division (−0.4% [95% PI: -2.7% to 2.1%]).

**Figure 2.**
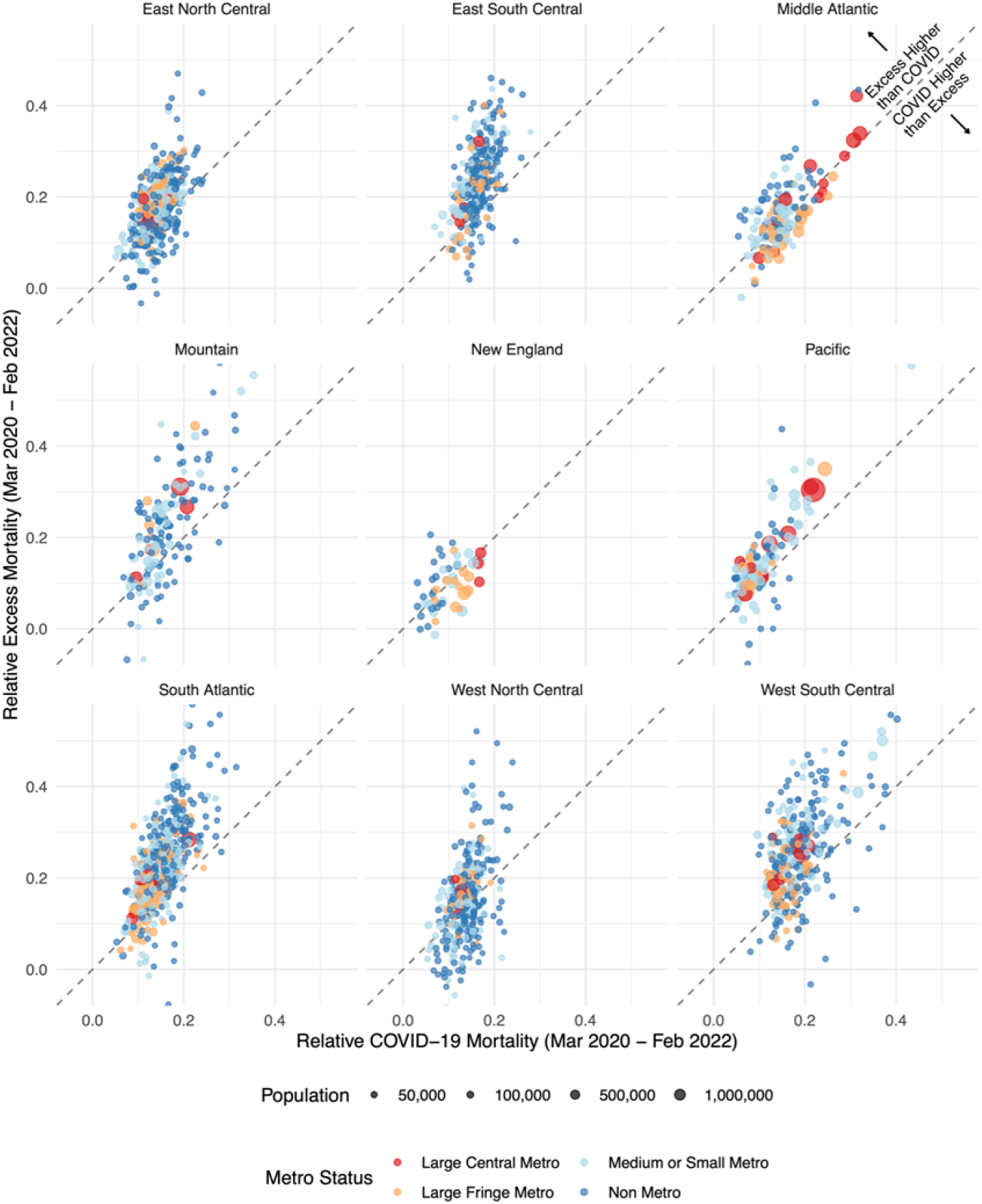
Comparison of excess deaths to COVID-19 deaths across U.S. counties by Census Division, March 2020 - February 2022 <u>Notes</u>: Each point in the graph represents a county and reflects its relative COVID-19 mortality from March 2020 to February 2022 (horizontal axis) and its relative excess mortality from March 2020 to February 2022 (vertical axis). Relative COVID-19 mortality is the ratio of COVID-19 deaths to expected deaths. We excluded counties with less than 10,000 residents to make the relationship between the two variables clearer. The 45 degrees line separates the plot into two parts. Points above the line saw more excess than COVID-19 mortality. Points falling below the line saw instead more COVID-19 mortality than excess mortality.

### Estimates Across Combinations of Metro-Nonmetro Categories and Census Divisions

In all 9 divisions, relative excess mortality for deaths not assigned to COVID-19 was higher in nonmetro counties than metro counties. The areas that had the highest relative excess mortality for deaths not assigned to COVID-19 were nonmetro counties in the South Atlantic division (12.9% [10.3% to 15.8%]), West South Central division (10.2% [95% PI: 7.6% to 13.1%]), Mountain division (9.5% [95% PI: 6.9% to 12.4%]), and East South Central division (8.8% [95% PI: 6.2% to 11.6%]). The areas that had negative relative excess mortality for deaths not assigned to COVID-19 were large metro counties in the New England division (−3.7% [95% PI: -6.0% to -1.2%]) and the Middle Atlantic division (−1.1% [95% PI: -3.4% to 1.4%]) and medium or small metro counties in the New England division (−2.2% [95% PI: -4.5% to 0.4%]) and the Middle Atlantic division (−0.1% [95% PI: -2.5% to 2.4%]). Nonmetro counties in these divisions had positive relative excess mortality for deaths not assigned to COVID-19.

### Temporal Patterns

**Figure 3** shows temporal variations in excess deaths, reported COVID-19 deaths, and excess deaths not assigned to COVID-19 by month across combinations of metro-nonmetro categories and Census divisions. Pronounced differences between excess deaths and reported COVID-19 deaths emerged early in the pandemic and persisted into the pandemic’s second year. Between March 2020 and February 2022, reported COVID-19 deaths tracked closely with monthly excess mortality estimates across all areas. Excess deaths not assigned to COVID-19 were typically more frequent during reported COVID-19 peaks than during the troughs between peaks. This is most apparent in areas that had the largest relative amounts of excess deaths not assigned to COVID-19 such as in the East South Central and South Atlantic divisions. In contrast, in the New England and Middle Atlantic divisions where COVID-19 deaths exceeded excess deaths in many areas, excess deaths not assigned to COVID-19 remained relatively stable over time, not changing during reported COVID-19 peaks. Finally, in areas where excess deaths not assigned to COVID-19 did rise with reported COVID-19 peaks, it appeared that the rise in excess deaths not assigned to COVID-19 often preceded or occurred concurrently with the rise in COVID-19 deaths.

**Figure 3.**
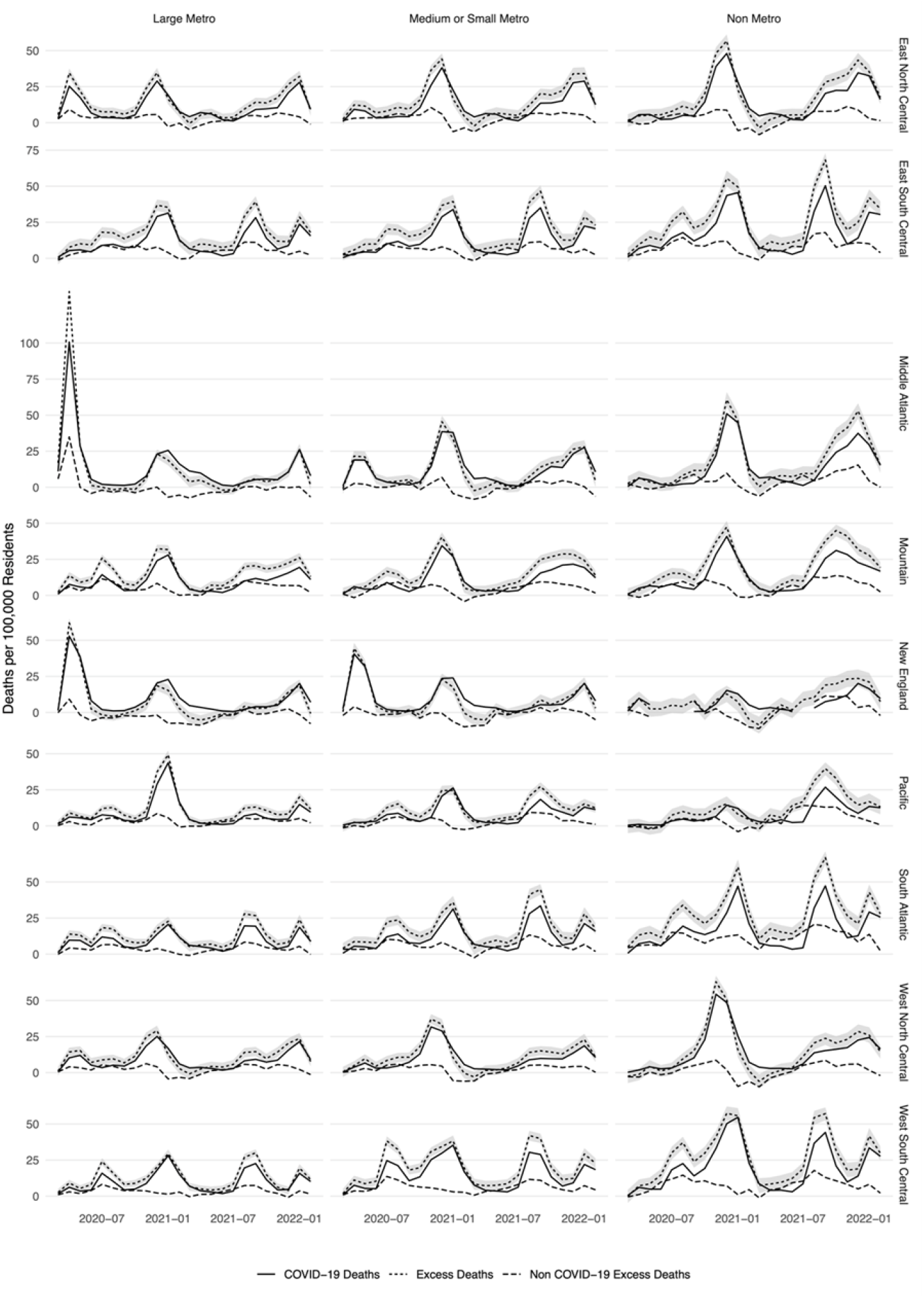
Monthly variation in COVID-19 and excess mortality across Census Divisions and metro/nonmetro categories

### Counties with the Largest Gaps Between Excess and COVID-19 Deaths

**Table 2** presents estimates of excess deaths not assigned to COVID-19 for the counties with the highest ratios of excess to COVID-19 deaths in each Census Division over the first two years of the pandemic. For example, Shelby, Tennessee had 2.4 times more excess deaths than COVID-19 deaths during the first two years of the pandemic. In absolute terms, this equaled 4,014 (95% PI: 3,562 to 4,517) excess deaths that were not assigned to COVID-19 during this period. Lafeyette, Louisiana had 2.5 times more excess deaths than COVID-19 deaths during the first two years of the pandemic. This equaled 654 (95% PI: 504 to 805) excess deaths not assigned to COVID-19 during this period. Estimates of the ratio and difference between excess and COVID-19 deaths for all 3,127 counties in the study are available in the **Supplementary Materials**.

**Table 2.** Excess mortality among U.S. counties with the highest excess to COVID-19 death ratios in each Census Division over the first two years of the pandemic.

| County | State | Excess/COVID | COVID-19 Deaths | Absolute Excess Mortality |  |  |  | Relative Excess Mortality |  |  |  |
| --- | --- | --- | --- | --- | --- | --- | --- | --- | --- | --- | --- |
|  |  |  |  | Total |  | Excluding COVID-19 |  | Total |  | Excluding COVID-19 |  |
|  |  |  |  | Median | PI (90%) | Median | PI (90%) | Median | PI (90%) | Median | PI (90%) |
| East North Central |  |  |  |  |  |  |  |  |  |  |  |
| Jackson | Illinois | 2.580 | 138 | 356 | ( 293 - 415) | 218 | ( 155 - 277) | 0.392 | ( 0.303 - 0.488) | 0.240 | ( 0.161 - 0.326) |
| Isabella | Michigan | 2.257 | 167 | 377 | ( 313 - 439) | 210 | ( 146 - 272) | 0.399 | ( 0.310 - 0.497) | 0.222 | ( 0.145 - 0.308) |
| Monroe | Wisconsin | 2.250 | 80 | 180 | ( 122 - 244) | 100 | ( 42 - 164) | 0.201 | ( 0.128 - 0.294) | 0.112 | ( 0.044 - 0.198) |
| Coles | Illinois | 2.165 | 124 | 268 | ( 203 - 333) | 144 | ( 79 - 209) | 0.276 | ( 0.197 - 0.367) | 0.149 | ( 0.077 - 0.230) |
| Scioto | Ohio | 2.062 | 257 | 530 | ( 425 - 636) | 273 | ( 168 - 379) | 0.252 | ( 0.193 - 0.319) | 0.130 | ( 0.076 - 0.190) |
| East South Central |  |  |  |  |  |  |  |  |  |  |  |
| Pontotoc | Mississippi | 2.618 | 110 | 288 | ( 237 - 341) | 178 | ( 127 - 231) | 0.458 | ( 0.350 - 0.592) | 0.283 | ( 0.188 - 0.401) |
| Shelby | Tennessee | 2.419 | 2,830 | 6,844 | ( 6,392 - 7,347) | 4,014 | ( 3,562 - 4,517) | 0.401 | ( 0.365 - 0.444) | 0.235 | ( 0.204 - 0.273) |
| Marshall | Mississippi | 2.372 | 137 | 325 | ( 264 - 381) | 188 | ( 127 - 244) | 0.399 | ( 0.301 - 0.502) | 0.231 | ( 0.145 - 0.321) |
| Hinds | Mississippi | 2.183 | 706 | 1,542 | ( 1,383 - 1,701) | 836 | ( 677 - 995) | 0.357 | ( 0.309 - 0.409) | 0.193 | ( 0.151 - 0.239) |
| Robertson | Tennessee | 2.153 | 209 | 450 | ( 365 - 529) | 241 | ( 156 - 320) | 0.302 | ( 0.231 - 0.374) | 0.162 | ( 0.099 - 0.226) |
| Middle Atlantic |  |  |  |  |  |  |  |  |  |  |  |
| Clinton | New York | 2.198 | 106 | 233 | ( 144 - 315) | 127 | ( 38 - 209) | 0.157 | ( 0.092 - 0.225) | 0.086 | ( 0.025 - 0.149) |
| Delaware | New York | 2.091 | 88 | 184 | ( 109 - 249) | 96 | ( 21 - 161) | 0.170 | ( 0.094 - 0.245) | 0.089 | ( 0.018 - 0.158) |
| Wayne | Pennsylvania | 1.947 | 150 | 292 | ( 213 - 369) | 142 | ( 63 - 219) | 0.231 | ( 0.159 - 0.311) | 0.112 | ( 0.048 - 0.184) |
| Tioga | New York | 1.916 | 107 | 205 | ( 138 - 268) | 98 | ( 31 - 161) | 0.211 | ( 0.134 - 0.295) | 0.101 | ( 0.031 - 0.177) |
| Franklin | New York | 1.903 | 72 | 137 | ( 75 - 201) | 65 | ( 3 - 129) | 0.149 | ( 0.077 - 0.236) | 0.071 | ( 0.003 - 0.151) |
| Mountain |  |  |  |  |  |  |  |  |  |  |  |
| Uintah | Utah | 2.717 | 53 | 144 | ( 93 - 190) | 91 | ( 40 - 137) | 0.279 | ( 0.164 - 0.403) | 0.176 | ( 0.070 - 0.291) |
| Douglas | Colorado | 2.111 | 416 | 878 | ( 729 - 999) | 462 | ( 313 - 583) | 0.280 | ( 0.222 - 0.332) | 0.148 | ( 0.095 - 0.194) |
| Douglas | Nevada | 2.084 | 107 | 223 | ( 150 - 288) | 116 | ( 43 - 181) | 0.202 | ( 0.128 - 0.277) | 0.105 | ( 0.037 - 0.174) |
| La Plata | Colorado | 2.069 | 87 | 180 | ( 127 - 240) | 93 | ( 40 - 153) | 0.252 | ( 0.165 - 0.367) | 0.130 | ( 0.052 - 0.234) |
| Sweetwater | Wyoming | 2.035 | 115 | 234 | ( 186 - 284) | 119 | ( 71 - 169) | 0.388 | ( 0.286 - 0.514) | 0.197 | ( 0.109 - 0.306) |
| New England |  |  |  |  |  |  |  |  |  |  |  |
| Washington | Maine | 4.198 | 43 | 180 | ( 116 - 241) | 138 | ( 73 - 198) | 0.204 | ( 0.122 - 0.292) | 0.155 | ( 0.077 - 0.240) |
| Washington | Vermont | 3.470 | 33 | 114 | ( 46 - 189) | 82 | ( 13 - 156) | 0.100 | ( 0.038 - 0.176) | 0.071 | ( 0.011 - 0.145) |
| Addison | Vermont | 3.095 | 21 | 65 | ( 15 - 112) | 44 | ( -6 - 91) | 0.098 | ( 0.021 - 0.182) | 0.066 | ( -0.008 - 0.147) |
| Bennington | Vermont | 2.714 | 21 | 57 | ( -9 - 126) | 36 | ( -30 - 105) | 0.057 | ( -0.009 - 0.136) | 0.036 | ( -0.028 - 0.113) |
| Rutland | Vermont | 2.550 | 40 | 102 | ( 17 - 183) | 62 | ( -22 - 143) | 0.069 | ( 0.011 - 0.130) | 0.042 | ( -0.014 - 0.102) |
| Pacific |  |  |  |  |  |  |  |  |  |  |  |
| Maui | Hawaii | 3.455 | 88 | 304 | ( 187 - 413) | 216 | ( 99 - 325) | 0.128 | ( 0.075 - 0.182) | 0.091 | ( 0.040 - 0.143) |
| Mendocino | California | 2.538 | 104 | 264 | ( 174 - 360) | 160 | ( 70 - 256) | 0.143 | ( 0.091 - 0.206) | 0.087 | ( 0.037 - 0.147) |
| Amador | California | 2.478 | 46 | 114 | ( 47 - 181) | 68 | ( 1 - 135) | 0.118 | ( 0.045 - 0.201) | 0.070 | ( 0.001 - 0.150) |
| Lane | Oregon | 2.401 | 421 | 1,011 | ( 756 - 1,261) | 590 | ( 335 - 840) | 0.131 | ( 0.095 - 0.169) | 0.076 | ( 0.042 - 0.112) |
| San Francisco | California | 2.392 | 704 | 1,684 | ( 1,341 - 2,055) | 980 | ( 637 - 1,351) | 0.147 | ( 0.114 - 0.185) | 0.085 | ( 0.054 - 0.122) |
| South Atlantic |  |  |  |  |  |  |  |  |  |  |  |
| Amherst | Virginia | 2.983 | 59 | 176 | ( 120 - 233) | 117 | ( 61 - 174) | 0.248 | ( 0.157 - 0.357) | 0.165 | ( 0.080 - 0.267) |
| Duplin | North Carolina | 2.903 | 185 | 537 | ( 471 - 597) | 352 | ( 286 - 412) | 0.573 | ( 0.470 - 0.681) | 0.376 | ( 0.285 - 0.470) |
| Clarendon | South Carolina | 2.658 | 152 | 404 | ( 347 - 459) | 252 | ( 195 - 307) | 0.529 | ( 0.424 - 0.648) | 0.330 | ( 0.239 - 0.433) |
| Halifax | North Carolina | 2.628 | 156 | 410 | ( 336 - 485) | 254 | ( 180 - 329) | 0.315 | ( 0.244 - 0.395) | 0.195 | ( 0.131 - 0.268) |
| Louisa | Virginia | 2.486 | 69 | 172 | ( 109 - 225) | 102 | ( 40 - 156) | 0.224 | ( 0.133 - 0.316) | 0.134 | ( 0.050 - 0.219) |
| West North Central |  |  |  |  |  |  |  |  |  |  |  |
| Beltrami | Minnesota | 2.625 | 88 | 231 | ( 170 - 289) | 143 | ( 82 - 201) | 0.275 | ( 0.188 - 0.369) | 0.170 | ( 0.091 - 0.257) |
| Warren | Missouri | 2.612 | 80 | 209 | ( 153 - 261) | 129 | ( 73 - 181) | 0.315 | ( 0.213 - 0.427) | 0.195 | ( 0.102 - 0.296) |
| Camden | Missouri | 2.336 | 149 | 348 | ( 283 - 412) | 199 | ( 134 - 263) | 0.353 | ( 0.269 - 0.447) | 0.202 | ( 0.127 - 0.285) |
| Cass | Minnesota | 2.220 | 50 | 111 | ( 59 - 160) | 61 | ( 9 - 110) | 0.171 | ( 0.084 - 0.266) | 0.094 | ( 0.013 - 0.183) |
| Itasca | Minnesota | 2.000 | 94 | 188 | ( 120 - 253) | 94 | ( 26 - 159) | 0.174 | ( 0.106 - 0.250) | 0.087 | ( 0.024 - 0.157) |
| West South Central |  |  |  |  |  |  |  |  |  |  |  |
| Iberville | Louisiana | 3.469 | 32 | 111 | ( 83 - 138) | 79 | ( 51 - 106) | 0.474 | ( 0.317 - 0.667) | 0.338 | ( 0.195 - 0.512) |
| Lafayette | Louisiana | 2.481 | 442 | 1,096 | ( 946 - 1,247) | 654 | ( 504 - 805) | 0.294 | ( 0.244 - 0.349) | 0.176 | ( 0.130 - 0.225) |
| Acadia | Louisiana | 2.241 | 245 | 549 | ( 470 - 626) | 304 | ( 225 - 381) | 0.439 | ( 0.354 - 0.533) | 0.243 | ( 0.170 - 0.325) |
| Webster | Louisiana | 2.213 | 167 | 370 | ( 304 - 437) | 202 | ( 137 - 270) | 0.366 | ( 0.283 - 0.464) | 0.201 | ( 0.127 - 0.287) |
| Vermilion | Louisiana | 2.193 | 181 | 397 | ( 330 - 467) | 216 | ( 149 - 286) | 0.345 | ( 0.271 - 0.432) | 0.188 | ( 0.122 - 0.265) |

**Notes:** Limited to counties with more than 30,000 residents and at least 20 COVID-19 deaths cumulatively over the period from March 2020 through February 2022.

## Discussion

In the present study, we generated estimates of excess mortality for counties across the U.S. and identified 268,176 excess deaths that were not assigned to COVID-19 during the first two years of the COVID-19 pandemic, which represented 23.7% of all excess deaths that occurred. Excess deaths were less likely to be assigned to COVID-19 in the Mountain division, in the South, and in nonmetro counties. The number of excess deaths exceeded COVID-19 deaths in all Census divisions except for the New England and Middle Atlantic divisions where there were more COVID-19 deaths than excess deaths in large metro areas and medium or small metro areas. In areas where substantial numbers of excess deaths were not assigned to COVID-19, rises in excess deaths not assigned to COVID-19 generally tracked temporally with rises in reported COVID-19 deaths. Contrary to prior literature which suggested that the majority of excess deaths not assigned to COVID-19 occurred early in the pandemic,^37^ differences between excess deaths and reported COVID-19 deaths were substantial in both the first and second year of the pandemic.

Although our study does not distinguish between unrecognized COVID-19 deaths and deaths indirectly related to the pandemic, emerging literature suggests that a large share of the excess deaths not assigned to COVID-19 represent unrecognized COVID-19 deaths.^12,13,38^ One study found that approximately 90% of excess mortality between March 2020 and April 2021 could be attributed to the direct effects of SARS-CoV-2 infection.^38^ Another study found significant temporal concordance between peaks in out-of-hospital COVID-19 deaths and excess deaths not assigned to COVID-19, with the latter preceding the former by 2 to 3 weeks.^14^ This suggests that many COVID-19 deaths go unrecognized during the beginning of COVID-19 surges when testing may be less frequent and medical providers are not expecting cases. Our study may provide further evidence to support this observation, as rises in excess deaths not assigned to COVID-19 typically preceded or occurred concurrently with rises in reported COVID-19 deaths in most Census divisions throughout the first two years of the pandemic. This possibility is also supported by investigative reporting during the pandemic which has documented widespread irregularities in cause-of-death assignment resulting from over-burdened and under-resourced death investigation systems.^16^ The individual county-level estimates generated in this study may be useful to reporters and public health officials interested in investigating possible irregularities in cause-of-death assignment within the death investigation system during the pandemic.

Discrepancies between COVID-19 and excess deaths are problematic because they have the potential to mislead scientists and policymakers about which areas were most heavily affected by the pandemic. Failure to accurately capture COVID-19 deaths also points to an urgent need to modernize the death investigation system in the United States, including expanding budgets for medical examiner officers and eliminating the coroner system.^15^ It can also affect individuals and families, as unrecognized COVID-19 deaths are not eligible for economic relief programs such as FEMA’s funeral assistance program.

Our study identified especially large discrepancies between COVID-19 and excess mortality rates in the Mountain division, in the South, and in nonmetro counties. More unrecognized COVID-19 deaths may have occurred in these areas as a result of limited COVID-19 testing and a greater share of deaths occuring outside of hospital settings, both of which complicate valid cause-of-death assignment.^8,14,39^ Another potential contributing factor is the greater reliance on coroners in these regions, who are typically elected and often hold other positions within the county such as sheriff-coroners.^40^ Medical training required for coroners is limited and highly variable across states,^41^ whereas medical examiners are physicians with extensive training in forensic pathology and death investigation.^42^ Coroners may also have more limited budgets for death investigation, affecting the likelihood they might pursue post-mortem COVID-19 testing.^15^ In fact, preliminary reporting has identified specific coroners across the United States who frequently defer to the decedent’s family regarding what they list as the underlying cause of death on the death certificate.^43^ Outside of COVID-19 mortality reporting, coroner systems have been shown to under-report opioid-related overdoses, and serious issues have been raised about the failures of medicolegal death investigation systems to monitor deaths in police custody.^44,45^

In the New England and Middle Atlantic divisions, we observed a different pattern around assignment of COVID-19 deaths than in other areas. In these divisions, a large share of counties had higher COVID-19 than excess death rates. Several explanations may exist for this pattern including that other causes of death (i.e. influenza) declined in these areas or that the economically privileged status of some of these counties shielded their residents from the negative indirect effect of the pandemic by allowing them to work-from-home and avoid household crowding. Finally, it is possible that deaths were over-assigned to COVID-19 in these areas due to different cause-of-death assignment protocols for COVID-19. For example, until March 2022, COVID-19 deaths in the state of Massachusetts included any death that occurred within 60 days of a COVID-19 diagnosis, which differed from other states and guidelines from the Council of State and Territorial Epidemiologists that recommended states use a 30 day window.^46^

Our study had several limitations. First, we were unable to distinguish between excess deaths that represented unrecognized COVID-19 deaths and excess deaths indirectly related to the pandemic. Future research should examine this distinction to clarify the extent to which the excess deaths not assigned to COVID-19 reported in this study represent under-reporting of COVID-19 deaths. Second, we used underlying cause of death data to identify deaths assigned to COVID-19 and thus did not identify deaths where COVID-19 was listed as a contributing cause or appeared elsewhere on the death certificate. Third, some counties and states have experienced prolonged reporting delays of COVID-19 deaths, which could affect our estimates of the proportion of excess deaths assigned to COVID-19, particularly in more recent months.

## Conclusion

In this study, we used death certificate data to compare excess deaths during the COVID-19 pandemic to reported COVID-19 deaths in counties across the U.S.. We found that many excess deaths were not assigned to COVID-19, and that contrary to prior reports, the number of excess deaths not assigned to COVID-19 was substantial in year two of the pandemic along with year one. Specific regions – the Mountain division, the South, and nonmetro areas – were identified as being less likely to report excess deaths as COVID-19 deaths, suggesting there may be a greater proportion of unrecognized COVID-19 deaths in communities in these areas. In contrast, many counties in the New England and Middle Atlantic Divisions reported more COVID-19 deaths than excess deaths. Overall, the extent to which reported COVID-19 deaths reflected the true mortality impact of the pandemic varied by time and place throughout the U.S. during the first two years of the pandemic. Moving forward, efforts to target resources to the communities most impacted by the pandemic should consider this variation.

## Data Availability

Data used in the study are publicly available from the US Centers for Disease Control and Prevention and US Census Bureau.

https://github.com/eugeniopaglino/county-month-excess

## Data and Materials Availability

Data used in the study are publicly available from the US Centers for Disease Control and Prevention and US Census Bureau. Additional details about the data and programming code for replication can be accessed at the linked GitHub repository: https://github.com/eugeniopaglino/county-month-excess.

## Supplementary Materials for

### Supplementary Text

#### Data Extraction from CDC WONDER

The CDC WONDER online database query system found at https://wonder.cdc.gov/ was used to extract all mortality data used in this project. To obtain death counts for all-causes mortality, we used the Multiple Cause of Death (Final) database from 1999-2020. We obtained two main sets of extracts, one for data at the county-month level and one for data at the county-year level (to investigate suppression). In order to minimize suppression, additional extracts were obtained at the wave level and the pandemic year level (pooling different months) for all figures and tables using these longer time periods. For all cause mortality extracts, the data request was submitted for the time period of interest using the request form with the following settings:

- Tab 1. Organize table layout: Group results by County and by: Month for monthly data, Year for yearly data, and no additional group for wave and pandemic year data
- Tab 4. Select time period of death: specific period
- Tab 6. Select underlying cause of death: *All* (All Causes of Death)
- Tab 8. Other options: checking Export Results. The request generates a text file.

To extract data on all causes of death for the time periods of March 2020 to February 2022, we used the Multiple Cause of Death (Provisional) database from 2018 – Last Month database. The data requests were submitted for each time period of interest using the request form with the following settings:

- Tab 1. Organize table layout: Group results by County and by: Month for monthly data, Year for yearly data, and no additional group for wave and pandemic year data
- Tab 4. Select time period of death: March 2020 to February 2022
- Tab 6. Select underlying cause of death: *All* (All Causes of Death)
- Tab 8. Other options: checking Export Results. The request generates a text file.

To extract counts of COVID deaths for March 2020 to February 2022 we used the Multiple Cause of Death (Provisional) database from 2018 – Last Month database. The data requests were submitted for each time period of interest using the request form with the following settings changed from the default:

- Tab 1. Organize table layout: Group results by County and by: Month for monthly data, Year for yearly data, and no additional group for wave and pandemic year data
- Tab 4. Select time period of death: March 2020 to February 2022
- Tab 6. Select underlying cause of death: U07.1 (COVID-19)
- Tab 8. Other options: checking Export Results. The request generates a text file.

#### Geographic Classifications

##### USDA/ERS/NCHS Metropolitan-Nonmetropolitan Categories

- **Large metros:** counties in metropolitan statistical areas with a population of more than 1 million (large central metros) and counties that surround the large central metros (large fringe metros).
- **Small or medium metros**: counties in metropolitan statistical areas with a population between 50,000 and 999,999.
- **Nonmetropolitan areas**: all other counties.

##### US Census Divisions

- **New England**: Connecticut, Maine, Massachusetts, New Hampshire, Rhode Island, Vermont
- **Middle Atlantic**: New Jersey, New York, Pennsylvania
- **East North Central**: Indiana, Illinois, Michigan, Ohio, Wisconsin
- **West North Central**: Iowa, Kansas, Minnesota, Missouri, Nebraska, North Dakota, South Dakota
- **South Atlantic**: Delaware, District of Columbia, Florida, Georgia, Maryland, North Carolina, South Carolina, Virginia, West Virginia
- **East South Central**: Alabama, Kentucky, Mississippi, Tennessee
- **West South Central**: Arkansas, Louisiana, Oklahoma, Texas
- **Mountain**: Arizona, Colorado, Idaho, New Mexico, Montana, Utah, Nevada, Wyoming
- **Pacific**: Alaska, California, Hawaii, Oregon, and Washington

**Supplementary Table.** Excess mortality across U.S. states over the first two years of the pandemic.

| State | COVID-19 Deaths | Excess/COVID | Absolute Excess Mortality |  |  |  | Relative Excess Mortality |  |  |  |
| --- | --- | --- | --- | --- | --- | --- | --- | --- | --- | --- |
|  |  |  | Total |  | Excluding COVID-19 |  | Total |  | Excluding COVID-19 |  |
|  |  |  | Median | PI (90%) | Median | PI (90%) | Median | PI (90%) | Median | PI (90%) |
| Alabama | 18,278 | 1.344 | 24,566 | ( 21,868 - 27,464) | 6,288 | ( 3,590 - 9,186) | 0.219 | (0.190 - 0.251) | 0.056 | ( 0.031 - 0.084) |
| Alaska | 1,118 | 1.969 | 2,202 | (1,884 - 2,498) | 1,084 | ( 766 - 1,380) | 0.238 | (0.197 - 0.279) | 0.117 | ( 0.080 - 0.154) |
| Arizona | 23,931 | 1.637 | 39,168 | (36,247 - 42,124) | 15,238 | (12,316 - 18,193) | 0.322 | (0.291 - 0.354) | 0.125 | ( 0.099 - 0.153) |
| Arkansas | 9,524 | 1.423 | 13,548 | (11,939 - 15,230) | 4,024 | ( 2,415 - 5,706) | 0.204 | (0.176 - 0.236) | 0.061 | ( 0.036 - 0.088) |
| California | 85,730 | 1.372 | 117,611 | (104,574 - 131,242) | 31,881 | (18,844 - 45,512) | 0.214 | (0.186 - 0.245) | 0.058 | ( 0.034 - 0.085) |
| Colorado | 10,341 | 1.568 | 16,212 | (14,141 - 18,363) | 5,871 | ( 3,800 - 8,022) | 0.201 | (0.171 - 0.234) | 0.073 | ( 0.046 - 0.102) |
| Connecticut | 9,673 | 0.920 | 8,902 | ( 7,294 - 10,626) | -771 | (-2,378 - 953) | 0.138 | (0.111 - 0.170) | -0.012 | (-0.036 - 0.015) |
| Delaware | 2,526 | 1.378 | 3,480 | ( 2,964 - 4,025) | 954 | ( 438 - 1,499) | 0.178 | (0.148 - 0.212) | 0.049 | ( 0.022 - 0.079) |
| District of Columbia | 1,428 | 1.527 | 2,181 | (1,886 - 2,518) | 753 | ( 458 - 1,090) | 0.218 | (0.183 - 0.261) | 0.075 | ( 0.045 - 0.113) |
| Florida | 60,319 | 1.323 | 79,787 | (69,603 - 90,311) | 19,468 | ( 9,284 - 29,992) | 0.185 | (0.158 - 0.215) | 0.045 | ( 0.021 - 0.071) |
| Georgia | 27,511 | 1.612 | 44,361 | (40,195 - 48,768) | 16,850 | (12,684 - 21,257) | 0.252 | (0.223 - 0.284) | 0.096 | ( 0.070 - 0.124) |
| Hawaii | 1,240 | 1.337 | 1,658 | (1,072 - 2,308) | 418 | ( -167 - 1,068) | 0.070 | (0.044 - 0.101) | 0.018 | (-0.007 - 0.047) |
| Idaho | 4,033 | 1.134 | 4,572 | ( 3,753 - 5,363) | 540 | ( -279 - 1,330) | 0.147 | (0.118 - 0.177) | 0.017 | (-0.009 - 0.044) |
| Illinois | 31,012 | 1.336 | 41,422 | (36,380 - 47,036) | 10,410 | ( 5,368 - 16,024) | 0.187 | (0.160 - 0.217) | 0.047 | ( 0.024 - 0.074) |
| Indiana | 19,985 | 1.222 | 24,417 | (21,208 - 27,781) | 4,432 | (1,223 - 7,796) | 0.180 | (0.153 - 0.210) | 0.033 | ( 0.009 - 0.059) |
| Iowa | 8,261 | 1.016 | 8,391 | ( 6,793 - 10,008) | 130 | (-1,467 - 1,747) | 0.134 | (0.105 - 0.163) | 0.002 | (-0.023 - 0.029) |
| Kansas | 7,411 | 1.291 | 9,566 | ( 8,232 - 11,068) | 2,156 | ( 821 - 3,657) | 0.171 | (0.144 - 0.203) | 0.038 | ( 0.014 - 0.067) |
| Kentucky | 12,861 | 1.485 | 19,094 | (16,702 - 21,487) | 6,233 | ( 3,841 - 8,626) | 0.191 | (0.163 - 0.221) | 0.062 | ( 0.038 - 0.089) |
| Louisiana | 14,312 | 1.717 | 24,571 | (22,298 - 26,860) | 10,259 | ( 7,986 - 12,548) | 0.267 | (0.236 - 0.299) | 0.111 | ( 0.085 - 0.140) |
| Maine | 2,133 | 1.286 | 2,743 | (1,994 - 3,584) | 610 | ( -138 - 1,451) | 0.089 | (0.063 - 0.120) | 0.020 | (-0.004 - 0.049) |
| Maryland | 13,182 | 1.284 | 16,920 | (14,369 - 19,659) | 3,738 | ( 1,187 - 6,477) | 0.163 | (0.135 - 0.195) | 0.036 | ( 0.011 - 0.064) |
| Massachusetts | 15,978 | 0.652 | 10,420 | ( 7,495 - 13,424) | -5,558 | (-8,482 - -2,553) | 0.085 | (0.060 - 0.112) | -0.045 | (-0.067 - -0.021) |
| Michigan | 28,198 | 1.313 | 37,026 | (32,192 - 42,125) | 8,828 | ( 3,994 - 13,927) | 0.183 | (0.156 - 0.214) | 0.044 | ( 0.019 - 0.071) |
| Minnesota | 10,383 | 1.214 | 12,608 | (10,302 - 14,977) | 2,226 | ( -80 - 4,594) | 0.136 | (0.108 - 0.165) | 0.024 | (-0.001 - 0.051) |
| Mississippi | 10,905 | 1.651 | 18,006 | (16,379 - 19,695) | 7,100 | ( 5,474 - 8,790) | 0.275 | (0.245 - 0.309) | 0.109 | ( 0.082 - 0.138) |
| Missouri | 16,839 | 1.413 | 23,794 | (20,811 - 27,023) | 6,955 | ( 3,972 - 10,184) | 0.187 | (0.160 - 0.218) | 0.055 | ( 0.031 - 0.082) |
| Montana | 2,972 | 1.206 | 3,585 | ( 2,995 - 4,152) | 613 | ( 23 - 1,180) | 0.167 | (0.135 - 0.198) | 0.028 | ( 0.001 - 0.056) |
| Nebraska | 3,698 | 1.085 | 4,013 | ( 3,101 - 4,957) | 315 | ( -596 - 1,259) | 0.114 | (0.086 - 0.145) | 0.009 | (-0.017 - 0.037) |
| Nevada | 9,698 | 1.279 | 12,399 | (11,122 - 13,792) | 2,701 | ( 1,424 - 4,094) | 0.234 | (0.205 - 0.267) | 0.051 | ( 0.026 - 0.079) |
| New Hampshire | 2,191 | 0.988 | 2,166 | (1,490 - 2,889) | -26 | ( -700 - 698) | 0.082 | (0.055 - 0.113) | -0.001 | (-0.026 - 0.027) |
| New Jersey | 28,111 | 0.872 | 24,512 | (20,666 - 28,404) | -3,598 | (-7,444 - 293) | 0.154 | (0.127 - 0.183) | -0.023 | (-0.046 - 0.002) |
| New Mexico | 7,280 | 1.605 | 11,682 | (10,702 - 12,728) | 4,402 | ( 3,422 - 5,448) | 0.302 | (0.269 - 0.338) | 0.114 | ( 0.086 - 0.145) |
| New York | 63,779 | 1.041 | 66,386 | (58,703 - 74,611) | 2,607 | (-5,075 - 10,832) | 0.204 | (0.176 - 0.235) | 0.008 | (-0.015 - 0.034) |
| North Carolina | 25,194 | 1.526 | 38,456 | (33,826 - 43,299) | 13,262 | ( 8,632 - 18,105) | 0.199 | (0.171 - 0.229) | 0.068 | ( 0.044 - 0.096) |
| North Dakota | 1,829 | 1.038 | 1,899 | (1,508 - 2,287) | 70 | ( -320 - 458) | 0.142 | (0.110 - 0.176) | 0.005 | (-0.023 - 0.035) |
| Ohio | 37,932 | 1.211 | 45,930 | (39,741 - 52,123) | 7,998 | ( 1,809 - 14,191) | 0.182 | (0.154 - 0.211) | 0.032 | ( 0.007 - 0.058) |
| Oklahoma | 13,652 | 1.321 | 18,031 | (15,992 - 20,245) | 4,379 | ( 2,340 - 6,593) | 0.217 | (0.188 - 0.251) | 0.053 | ( 0.028 - 0.082) |
| Oregon | 6,054 | 1.666 | 10,088 | ( 8,152 - 12,011) | 4,034 | ( 2,098 - 5,957) | 0.131 | (0.103 - 0.160) | 0.052 | ( 0.027 - 0.079) |
| Pennsylvania | 40,197 | 0.949 | 38,144 | (31,519 - 44,935) | -2,052 | (-8,677 - 4,738) | 0.137 | (0.110 - 0.165) | -0.007 | (-0.030 - 0.017) |
| Rhode Island | 2,918 | 0.637 | 1,859 | (1,274 - 2,464) | -1,059 | (-1,643 - -453) | 0.087 | (0.058 - 0.118) | -0.049 | (-0.075 - -0.022) |
| South Carolina | 16,622 | 1.557 | 25,886 | (23,381 - 28,555) | 9,264 | ( 6,759 - 11,933) | 0.249 | (0.220 - 0.282) | 0.089 | ( 0.063 - 0.118) |
| South Dakota | 2,235 | 1.210 | 2,704 | ( 2,225 - 3,155) | 469 | ( -9 - 920) | 0.163 | (0.130 - 0.196) | 0.028 | (-0.001 - 0.057) |
| Tennessee | 21,891 | 1.479 | 32,372 | (28,828 - 36,103) | 10,482 | ( 6,937 - 14,212) | 0.219 | (0.191 - 0.251) | 0.071 | ( 0.046 - 0.099) |
| Texas | 81,976 | 1.359 | 111,428 | (101,437 - 121,727) | 29,452 | (19,461 - 39,751) | 0.265 | (0.236 - 0.297) | 0.070 | ( 0.045 - 0.097) |
| Utah | 4,009 | 1.282 | 5,138 | (4,100 - 6,181) | 1,129 | ( 91 - 2,172) | 0.129 | (0.101 - 0.160) | 0.028 | ( 0.002 - 0.056) |
| Vermont | 454 | 1.905 | 865 | (465 - 1,226) | 411 | ( 11 - 772) | 0.068 | (0.036 - 0.100) | 0.033 | ( 0.001 - 0.063) |
| Virginia | 16,504 | 1.695 | 27,978 | (24,504 - 31,496) | 11,474 | ( 8,000 - 14,992) | 0.196 | (0.168 - 0.227) | 0.081 | ( 0.055 - 0.108) |
| Washington | 10,005 | 1.328 | 13,284 | (10,304 - 16,326) | 3,278 | ( 299 - 6,321) | 0.110 | (0.083 - 0.138) | 0.027 | ( 0.002 - 0.054) |
| West Virginia | 6,018 | 1.717 | 10,333 | ( 9,108 - 11,544) | 4,315 | ( 3,090 - 5,526) | 0.222 | (0.191 - 0.255) | 0.093 | ( 0.065 - 0.122) |
| Wisconsin | 12,214 | 1.273 | 15,546 | (12,862 - 18,373) | 3,332 | ( 648 - 6,159) | 0.141 | (0.114 - 0.171) | 0.030 | ( 0.006 - 0.057) |
| Wyoming | 1,642 | 1.668 | 2,739 | ( 2,446 - 3,051) | 1,097 | ( 804 - 1,409) | 0.273 | (0.237 - 0.314) | 0.109 | ( 0.078 - 0.145) |
| Total | 866,187 | 1.310 | 1,134,364 | (996,869 - 1,278,540) | 268,176 | (130,682 - 412,353) | 0.194 | (0.166 - 0.224) | 0.046 | ( 0.022 - 0.072) |

### Supplementary Data

County-month estimates of excess deaths, COVID-19 deaths, non-COVID-19 excess deaths, and the ratio of excess deaths to COVID-19 deaths are available at the linked GitHub repository: https://github.com/eugeniopaglino/county-month-excess.

